# The unedited acceptance rate: a review-fidelity metric for human-in-the-loop clinical AI

**DOI:** 10.64898/2026.11.19.26368546

**Authors:** Sofian Youssef, Omar Elalfy, Joseph Davids

**Author notes:** Corresponding author: Omar Elalfy, BSc, MBBCh, MPH, Wai Health Inc, Palo Alto, CA, United States.

## Abstract

**Objective:** Guidance for large language model (LLM) drafting in clinical care relies on human review, yet review is rarely measured as a control. We aimed to define computable measures of review fidelity, establish which a deployed system can compute, and pre-specify their first analysis.

**Materials and Methods:** For a multi-agent clinical system in routine use at one private London outpatient clinic, we defined acceptance coverage, attribution completeness, the unedited acceptance rate (UAR), a bounded normalized edit distance, availability latency and an unedited fast-approval rate (UFA). We checked each measure against the system’s code and database structure at a fixed commit on four approval routes and pre-specified a prospective 12-week analysis.

**Results:** The UAR equals the mass at zero of the edit-distance distribution, and availability latency bounds reading time from above only. Clinical documentation keeps draft and signed text with a recorded signer, so the core measures are computable there. Help-desk edit telemetry is written only when the agent’s version has been stored. The WhatsApp agent route records no edit signal, and its approver need not be named. No route records when a draft is opened. Documentation accept events were not written before a change committed on November 16, 2026. Model-generated content, including live voice-agent speech, also reaches patients without approval.

No measured values are reported.

**Discussion:** Acceptance alone cannot separate accurate drafts from approval without reading, and three of four routes cannot yet support the family.

**Conclusion:** Review fidelity can be specified as telemetry at the approval event. Validation against missed errors is required.

## Background and Significance

Human review is the safeguard on which current guidance for LLM drafting in clinical care depends. NHS England guidance on ambient scribing products requires users to review and approve outputs before further action is taken.^1^ The EU Artificial Intelligence Act requires human oversight of high-risk systems and asks that the people exercising it remain aware of automation bias.^2^ Medicines and Healthcare products Regulatory Agency (MHRA) guidance of July 2026 states that an ambient voice product that only transcribes or summarizes an encounter for clinician review is not a medical device, and its device example is an “AVT agent” that “finalises and saves transcripts to the electronic patient record without requiring a clinician’s review” and also autonomously determines follow-up tests and places “the relevant orders without clinician input or confirmation.“3 NHS England guidance, by contrast, states that generative summarization “likely would qualify as a medical device.”1 In each text the review step carries safety weight. None specifies how the quality of that review should be measured.

The evidence on deployed LLM tools mostly describes the tools. Randomized trials of ambient scribes measured documentation time and clinician well-being,^4,5^ and a multisite cohort measured time and visit volume.^6^ Studies of drafted replies to patient portal messages measured utilization, read time and reply time.^7,8^ A newer body of work examines what clinicians do to drafts. In 23,760 ambient notes, 84.4% were edited before sign-off, variation was explained more by the individual clinician than by specialty, and notes signed later were edited less intensively.^9^ A qualitative analysis found that edits concentrated on orders, symptoms, prescriptions and diagnoses rather than on style.^10^ In 14,350 AI-assisted portal replies, every edit category was associated with longer response time.^11^ A 1-year study used text similarity between draft and final reply as a measure of draft retention,^12^ a cohort of 983,154 responses separated adoption, use and habituation,^13^ an editing-load framework estimated how much a clinician would edit a given draft,^14^ and clinicians’ decisions to use drafted portal replies correlated with automated text metrics.^15^ These are research analyses of logs, samples or test sets. To our knowledge, none was reported as a monitored control within the deploying organization’s clinical safety process.

Unedited approvals carry particular safety weight, because an error in the draft then reaches the patient or the record unchanged. In a simulation study, 35% to 45% of erroneous drafts were submitted entirely unedited by primary care physicians,^16^ and assessors judged that 7.1% of LLM drafts in a radiation oncology messaging simulation posed a risk of severe harm if left unedited.^17^ Automation bias is established in clinical decision support and increases with workload, time pressure and verification complexity.^18–20^ It persists after training. In a randomized trial, physicians who had completed 20 hours of AI-literacy training had diagnostic accuracy 14.0 percentage points lower than controls when LLM suggestions contained deliberate errors.^21^ Pharmacists and trainees accepted 73.9% of incorrect and 80.2% of correct AI recommendations,^22^ which indicates that acceptance can be almost insensitive to correctness. Exposure to AI support may also reduce unaided performance.^23^

Regulators have begun to treat review quality as a quantity that changes over time. The MHRA AI Airlock phase 2 report concluded that human oversight may be inadequate to treat as a fixed safety control, observed that users applied less scrutiny as accurate output became familiar, and identified review time and edit depth as potential indirect indicators of engagement quality.^24^ The UK National Commission into the Regulation of AI in Healthcare asked that oversight design consider automation bias and that devices log performance to detect drift.^25^ Unedited acceptance has also been reported as a benefit. Trade press reported that 92% of AI-generated prior-authorization responses at one customer of an electronic health record vendor were accepted without edits.^26^ Without a measure of review, such a figure is compatible both with accurate drafts and with approval without reading. Commentators have called for accountable authorship and auditability of drafted messages.^27^

To our knowledge, no published deployment reports review fidelity as continuous telemetry computed at the point where a named person commits drafted content.

We considered that a usable measure of review fidelity should have five properties. It should be defined on a recorded approval event, be insensitive to formatting, be read jointly with measures that separate competing explanations, be protected from becoming a target or a tool for individual surveillance, and be analyzed under a plan fixed before the data exist. A sixth requirement follows from the first. A measure is usable only where the system actually records the event it is defined on, and that has to be checked in the code rather than assumed from the design.

## Objective

We aimed to define the unedited acceptance rate (UAR) family formally, to establish for each approval route of a multi-agent clinical system in routine outpatient use which measures its instruments can compute and which instruments do not yet exist, to set out an interpretation framework and proposed governance uses, and to pre-specify the analysis of a prospective 12-week measurement window.^28,29^ This article reports the properties of the measures and the state of the instruments, and no measured values.

## Materials and Methods

### Setting and system

The system studied, wai, is a multi-agent clinical system in routine use at a private outpatient clinic in London, developed and operated by Wai Health Inc.^28,29^ Version 0.2 of its action inventory (September 8, 2026), compiled from the code, lists 153 live agent actions in 23 functional domains.^28,30^ The inventory states that 29 of these actions reach a patient or the clinical record with no person accepting them. It does not itemize the 29. Its own list of such actions names 33 live rows, of which 30 are classed as having no acceptance step, one as partial acceptance, one as not applicable and one as an action that has since been converted to a draft. Each no-acceptance row has an entry in the clinic’s hazard log.^30^ Some of these actions carry model-generated content. They include the voice agent speaking live to a patient, a live recap pushed to the patient’s screen during a consultation, live translated captions, model-extracted details and model-written descriptions of patient media written to the record, and a command-chat tool that can change the body of a clinical document. Such content reaches patients or the record without approval and lies outside every measure defined here.

Drafted text that does pass through approval does so on four routes, which are the measurement routes of this paper. The first is the help desk, on which staff approve drafted replies to patient messages on several channels, including email and in-app messages. The second is the WhatsApp agent, whose drafts are approved in the administration console or from an approval card posted to the team’s Slack workspace. The third is clinical documentation, in which a clinician signs a drafted document. The fourth is approval cards for actions proposed by the operations agent in Slack, which offer approve or reject. Table 4 lists the models configured for drafting on the first three routes.

### Design and reporting

This is a methods study with a pre-specified analysis plan. Every statement about the system was checked against its code at commit 687f45e295 (November 19, 2026), against its safety documents in the same repository, and against read-only queries of database structure that returned no rows. Commit dates are reported as such and are not deployment dates. Reporting follows published guidance for analyses of metadata on electronic health record use^33^ and the TRIPOD-LLM items that apply to a methods study of LLM use in a health care setting,^34^ with a completed checklist in Supplementary File 1. The full analysis plan is given in Supplementary File 2, in the fields of the Open Science Framework (OSF) Preregistration template. It will be registered on the OSF with a public time stamp before the measurement window opens, so that no data it governs exist when it is fixed.

### Measurement surface

Review fidelity is measurable only where review is a recorded event. For an approval event i, the measures need five things: the drafted text Di, the committed text Ci, the identity of the approver, the time tavail,i at which the draft became available for review and the commit time tcommit,i (Figure 1). A sixth, the time the draft was first opened for review, would allow a tighter latency measure. Whether each route records each of these is reported in Results and Table 3.

**Figure 1.**
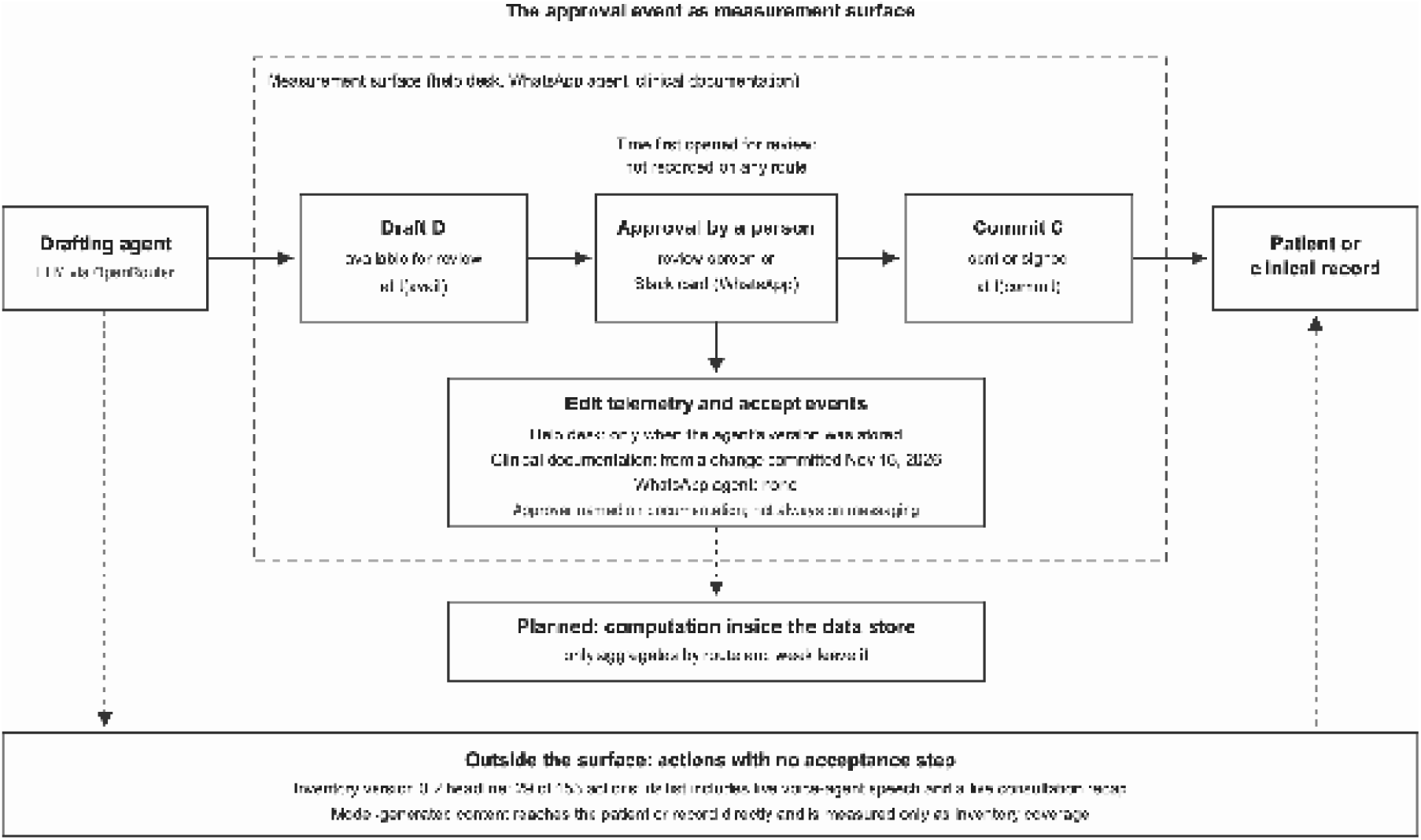
The approval event as measurement surface. A drafting agent writes a draft, which becomes available for review at its availability time. A person approves it, and the committed text takes effect at the commit time. Edit telemetry is written on the help desk only when the review screen has stored the agent’s version, on clinical documentation from a change committed on November 16, 2026, and not on the WhatsApp agent route. No route records when a draft is first opened. Under the analysis plan, distances, latencies and reading floors are computed inside the clinical data store and only aggregates by route and week leave it. Inventoried actions that take effect without an acceptance step, including live voice-agent speech, lie outside the surface.

### Metric definitions

Let Ar,w be the set of approval events on route r whose commit time falls in week w and which pass the completeness rule of the analysis plan. Table 1 gives each measure with its numerator, denominator and missingness rule.

**Table 1.** Definitions of the review-fidelity measures, with numerators, denominators and missingness rules. All measures are computed by route and week on normalized text.

| Measure | Definition | Numerator | Denominator | Reported as | Missingness rule |
|---|---|---|---|---|---|
| Acceptance coverage (AC) | Share of commits that pass through a captured approval | Commits matched to an approval event with complete capture | Commits in the route's commit record | n/N, % (Wilson 95% CI) | Commits with no matching approval event counted as uncovered |
| Attribution completeness | Share of approval events naming a person as approver | Events whose approver field identifies a named person | All approval events | n/N, % (Wilson 95% CI) | Blank, surface-name and generic labels counted as not attributed |
| Inventory coverage | Inventoried actions that take effect without human acceptance | Actions with no acceptance step, from an itemized list | All inventoried actions | n of N | Not applicable (document-derived) |
| Unedited acceptance rate (UAR) | Share of approvals committing the draft unchanged after normalization | Events with δ = 0 | Complete approval events | n/N, % (Wilson 95% CI) | Incomplete events excluded and counted |
| Edit depth | Distribution of δ among edited approvals | Not applicable | Complete events with δ > 0 | Median, 75th and 90th percentiles | As for UAR |
| Availability latency (L) | Time from draft availability to commit | Not applicable | Complete events with both times | Median, 90th percentile | Missing latency reported separately |
| Dwell latency | Time from first opening for review to commit | Not applicable | Events with an open time | Not computable | No route records open events |
| Unedited fast-approval rate (UFA) | Share of approvals committed unedited faster than the reading floor f | Unedited events with L < f | Complete events with a latency value | n/N, % (Wilson 95% CI), with the conditional share among unedited events | Computed only on events with latency |
| Disposition | Share of drafts rejected or expired without commit | Rejected or expired drafts | Drafts produced | n/N, % | Reported only where discards are captured |

δ, Levenshtein distance in UTF-16 code units divided by the length of the longer normalized text; f, reading floor at 300 words per minute (sensitivity analysis 238 words per minute); CI, confidence interval.

#### Normalization

Before comparison, both texts are normalized by a function N that collapses every run of whitespace, including line breaks, to a single space and trims leading and trailing whitespace. For clinical documentation, N first removes markup tags and non-breaking-space entities. N applies no Unicode normalization and removes no appended elements. Removal is not needed, because the committed text is taken at the approval step, before translation or channel formatting is applied. This is the normalization that the system’s edit telemetry applies, and the analysis uses it unchanged so that stored and recomputed values agree.

#### Unedited acceptance rate

UARr,w is the share of events in Ar,w with N(Ci) = N(Di).

Normalized edit distance. δi = lev(N(Di), N(Ci)) / max(|N(Di)|, |N(Ci)|), where lev is the Levenshtein distance^35^ and |·| is length, both counted in UTF-16 code units, as implemented in the system. This differs from a count over Unicode code points only for characters outside the Basic Multilingual Plane, such as emoji. A word-level version computed over whitespace-delimited tokens is a pre-specified sensitivity analysis. The distribution of δi among edited approvals (δi > 0), described as edit depth, is reported separately from the UAR.

#### Approval latency

Availability latency is Li = tcommit,i − tavail,i, summarized by the median and 90th percentile.

Dwell latency, measured from the first opening of the draft for review, is defined for completeness but cannot be computed in this deployment, because no route records that event.

#### Unedited fast-approval rate

For each event, a reading floor fi = 60·ni/ρ seconds is defined, where ni is the word count of N(Di) and ρ is a reading rate of 300 words per minute, the upper end of the typical adult silent reading range for English non-fiction.^36^ A rate of 238 words per minute, the meta-analytic mean,^36^ is a sensitivity analysis. Among events with a latency value, UFAr,w is the share committed unedited with Li < fi.

Because this joint share moves with the UAR, the conditional share of unedited approvals that were fast is reported beside it. The UFA is the only measure in the family that can indicate approval without reading.

#### Acceptance coverage

ACr,w is the share of commits on route r in week w, counted from a record written separately from the edit telemetry, that are matched to an approval event with complete capture. The commit record is the communications audit for the help desk, the draft status for the WhatsApp agent route, and the signed document, with its signature record and audit-chain event, for clinical documentation, which does not pass through the communications audit.

#### Attribution completeness

The share of approval events whose approver field identifies a named person, as opposed to being blank, holding the name of the approval surface, or holding a generic label. It is reported because the disclosure rules and any reading of a “named approver” depend on it.

#### Inventory coverage

At inventory level, coverage is the number of inventoried actions that take effect without human acceptance.

#### Disposition

Where discards are recorded, the share of drafts rejected or expired without commit.

### Interpretation framework

The measures are read jointly, by route and over time, under five principles. First, no single measure is interpreted alone. A high UAR is consistent with an accurate model on routine content and with approval without reading, and a low UAR is consistent with attentive review and with a poor model. Second, an unedited approval is not evidence that review was absent, and an edit is not evidence that review was adequate. A clinician can verify a correct draft without changing it, and a superficial edit does not show that the content was checked. The measures bound behavior and do not observe cognition, which is why a calibration audit is part of the method.

Third, latency is informative in one direction only. Availability latency includes queue time and is therefore an upper bound on reading time, so a latency below the reading floor is evidence against complete reading, whereas a long latency is not evidence for it. This holds wherever a draft is read, because no draft can be read before it exists. A dwell latency would not have the same property here, because WhatsApp agent drafts can be read, and approved, from a Slack card outside the review screen. Fourth, case mix drives every measure, so readings are stratified by route, draft type and, where it is recorded, model version, and pooled figures describe composition only. Fifth, instrument failure is ruled out before behavior is interpreted. A reading that meets a pre-specified failure condition is reported as an instrument finding. Table 2 sets out the joint readings.

**Table 2.** Joint interpretation of the measures within one route and stratum, after instrument-failure conditions have been excluded.

| Pattern | Readings consistent with the pattern | Next step |
|---|---|---|
| High UFA | Approval without reading | Examine route workflow and workload; calibration audit of fast unedited approvals |
| High UAR, low UFA, stable edit depth | Accurate drafts on routine content, or slow approval without checking | Calibration audit to estimate missed errors among unedited approvals |
| Low UAR, high edit depth | Attentive review, or a draft quality problem | Review model and prompt version; edit categories |
| Low UAR, low edit depth | Cosmetic or stylistic editing, possibly target-seeking | Edit categories; check for Goodhart effects |
| UAR rising and latency falling after a model change | Better drafts, or habituation and reduced scrutiny | Change assessment; calibration audit before and after the change |
| UAR of exactly 0 or 1 with many events | Capture defect until shown otherwise | Apply instrument-failure rule; verify both capture arms |
| Low attribution completeness | Approval through a shared surface or an unauthenticated session | Report at route level only; treat as a hazard, not a behavior |

UAR, unedited acceptance rate; UFA, unedited fast-approval rate.

### Proposed governance uses

The plan proposes three uses, none of which is yet in operation. The first is threshold monitoring. On each route, a sustained rise in UFA, or a sustained rise in UAR together with falling latency, would trigger examination of that route’s approval practice by the clinical safety officer. The trigger would start a human review and impose no automated sanction. The second is change evaluation. Every model or prompt change would be handled as a clinical change, with the UAR, edit depth, latency and UFA compared across it on each route. A version whose drafts are accepted with materially less review should be assessed with more caution, irrespective of its benchmark performance. The third is a calibration audit, in which a second clinician reviews a stratified sample of approved drafts (unedited and edited, within and below the reading floor) for content the approval should have corrected. The audit is the link between the telemetry and missed errors. Its sample size, its definition of a missed error and the blinding of the second clinician to stratum have not been fixed, and the audit design will be registered separately before its first sample is drawn. The hazard log already records the underlying gap. Its entry HZ-65, “We cannot tell a real acceptance from a reflexive one”, was open and marked urgent in version 0.6 (committed November 9, 2026), which notes that the help-desk and WhatsApp surfaces are still not measured.

Any measure used to steer behavior is exposed to Goodhart effects, in which the measure ceases to track the property it was chosen to represent once it becomes a target.^37^ The mitigations are structural. The measures are never reported as individual performance scores and never feed performance management, because the purpose is to detect systemic approval without reading and an individual target would reward cosmetic edits. Triggers lead to examination of a route’s workflow, not of a person. The calibration audit assesses content directly and cannot be satisfied by editing. Staff who approve drafts will be told of the measures and their purpose before the window opens, and the date will be recorded in an amendment to the registration.

### Pre-specified analysis plan

The plan is summarized here and given in full in Supplementary File 2.

#### Window

Twelve consecutive calendar weeks (Monday to Sunday, Europe/London), starting at 00:00 on the first Monday at least 14 days after the public time stamp of the OSF registration. The window is prospective, so no data it covers exist when the plan is registered.

#### Instruments and change points

Before the window opens, an amendment will record the configured models and the deployment date of any change made after commit 687f45e295 to the instruments in Table 3. A measure is computed on a route only from the deployment date of the instrument it depends on. A change to an instrument or a model during the window is a change point, and estimates are reported on either side of it.

**Table 3.**
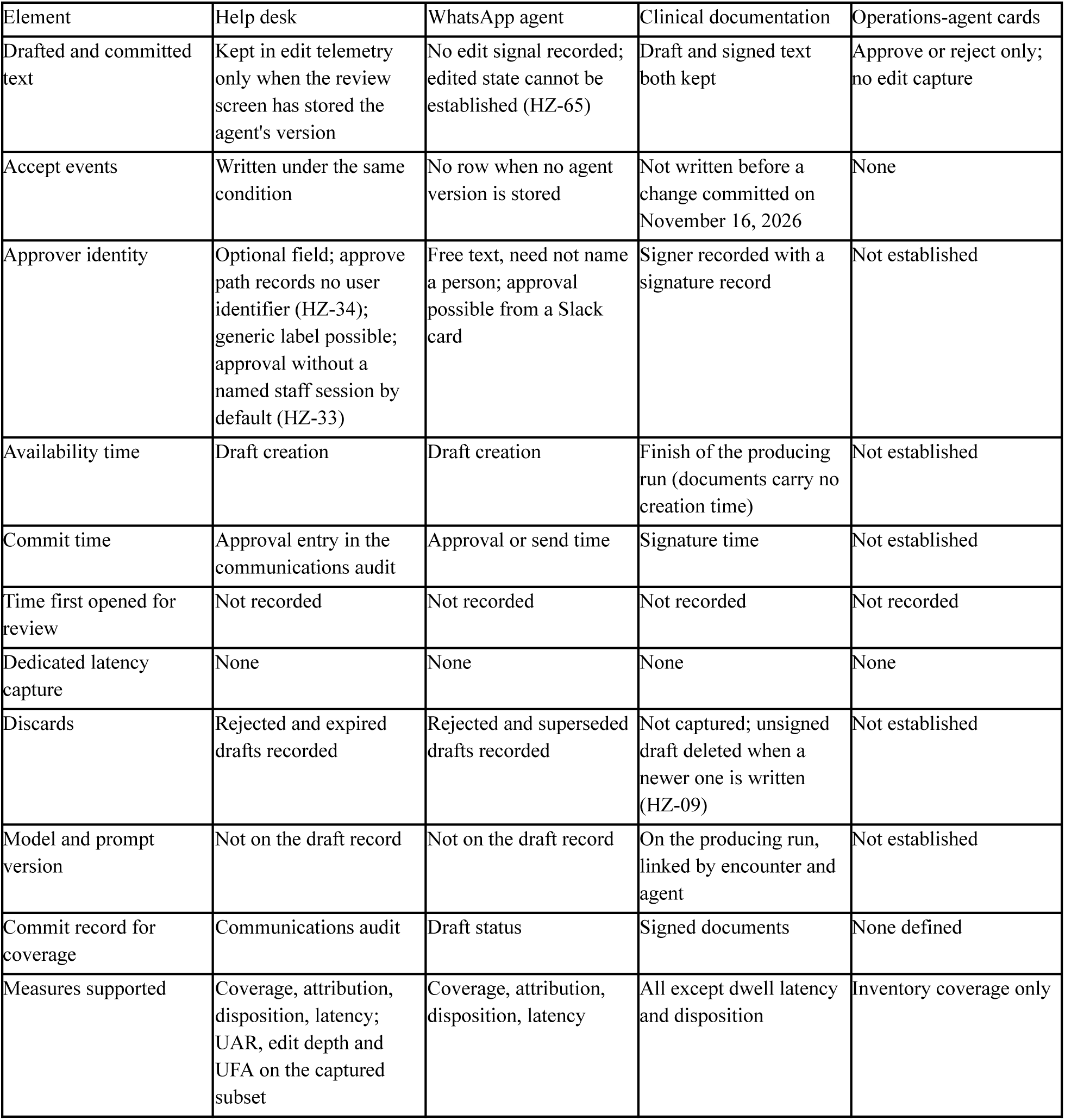
What each approval route records at commit 687f45e295 (November 19, 2026), and the measures it supports. Hazard identifiers refer to the clinic’s hazard log, version 0.6.

| Element | Help desk | WhatsApp agent | Clinical documentation | Operations-agent cards |
|---|---|---|---|---|
| Drafted and committed text | Kept in edit telemetry only when the review screen has stored the agent's version | No edit signal recorded; edited state cannot be established (HZ-65) | Draft and signed text both kept | Approve or reject only; no edit capture |
| Accept events | Written under the same condition | No row when no agent version is stored | Not written before a change committed on November 16, 2026 | None |
| Approver identity | Optional field; approve path records no user identifier (HZ-34); generic label possible; approval without a named staff session by default (HZ-33) | Free text, need not name a person; approval possible from a Slack card | Signer recorded with a signature record | Not established |
| Availability time | Draft creation | Draft creation | Finish of the producing run (documents carry no creation time) | Not established |
| Commit time | Approval entry in the communications audit | Approval or send time | Signature time | Not established |
| Time first opened for review | Not recorded | Not recorded | Not recorded | Not recorded |
| Dedicated latency capture | None | None | None | None |
| Discards | Rejected and expired drafts recorded | Rejected and superseded drafts recorded | Not captured; unsigned draft deleted when a newer one is written (HZ-09) | Not established |
| Model and prompt version | Not on the draft record | Not on the draft record | On the producing run, linked by encounter and agent | Not established |
| Commit record for coverage | Communications audit | Draft status | Signed documents | None defined |
| Measures supported | Coverage, attribution, disposition, latency; UAR, edit depth and UFA on the captured subset | Coverage, attribution, disposition, latency | All except dwell latency and disposition | Inventory coverage only |

**Table 4.** Models configured for drafting on the measured routes, as read from code and live service configuration on November 19, 2026. All are reached through the OpenRouter routing service.

| Route | Model chain (provider) | Notes |
|---|---|---|
| Clinical documentation (note engine) | deepseek/deepseek-v4-pro (DeepSeek); fallbacks deepseek/deepseek-v4-pro, then z-ai/glm-5.2 (Z.ai) | Set by service configuration; if the setting is absent the code falls back to deepseek/deepseek-v4-flash. When the live value was set is not recorded |
| Help desk drafting | deepseek/deepseek-v4-pro (DeepSeek), then moonshotai/kimi-k3 (Moonshot AI), then deepseek/deepseek-v4-pro | Code defaults; no override is set. Reasoning effort medium by default from a change committed on October 8, 2026 |
| WhatsApp agent drafting | As for the help desk | As for the help desk |

Draft records on the help-desk and WhatsApp agent routes do not record which model in the chain answered.
Training cut-off dates are not reported. The configuration in force when the window opens will be recorded in an amendment to the registration.

#### Unit, eligibility and completeness

The unit of analysis is the approval event. The system has no field that marks test records, so events on records in a list of test records fixed before the window opens are excluded. The operations-agent route is excluded from all measures except inventory coverage. An event is complete if it has a drafted text, a committed text or a stored unedited flag, an approver field and a commit time. Incomplete events are excluded from the UAR and edit depth and reported as a completeness proportion by route and week. The UFA is computed only on events with a latency value.

#### Estimators and precision

Proportions are reported as counts with Wilson score 95% confidence intervals,^38^ edit depth as the median, 75th and 90th percentiles of δ among edited approvals, and latency as the median and 90th percentile, by route and week. Approvals cluster within approvers and weeks, so Wilson intervals are descriptive, and a cluster bootstrap over weeks (2,000 replicates) is a sensitivity analysis for window-level estimates, which are interpreted as approximate. The half-width of a 95% Wilson interval near 0.5 is about 0.20 at 20 events, 0.10 at 96 and 0.05 at 384, so weekly estimates on low-volume routes are shown as series and not compared week by week. No trend test is pre-specified, and any trend statement will be labeled exploratory. Strata are route, draft type, tertile of draft word count within route and, for clinical documentation only, model and prompt version.

#### Disclosure control

Cells with fewer than five events are suppressed, with secondary suppression so that no suppressed value can be derived by subtraction. Cells whose approvals come from fewer than three distinct named approvers are reported only after further aggregation. Events without a named approver are reported only at route level over the whole window. Counts by approver are never reported.

#### Instrument-failure conditions

A route-week is an instrument failure if (1) acceptance coverage is below 95%; (2) completeness among captured events is below 95%; (3) the UAR is exactly 0 or 1 with at least 20 events, unless both capture arms are shown to work on synthetic records in a non-production environment; or (4) for clinical documentation, more than 5% of events cannot be linked to their producing run. Failed route-weeks are counted, reported as instrument findings and excluded from behavioral interpretation. Condition 3 triggers verification rather than classifying behavior, because on a route whose true unedited share is 0.95, 20 approvals that are all unedited occur with probability 0.36.

Version control. Extraction queries will compute distances, latencies and reading floors inside the clinical database and export only aggregates by route, stratum and week. Their identifiers will be recorded in an amendment before extraction, and every reported aggregate will cite the query version and extraction date. Deviations will be reported with reasons.

## Results

### Properties of the measures

Four properties follow directly from the definitions. They fix how the measures are interpreted and are not empirical findings. First, δi = 0 if and only if N(Ci) = N(Di), so the UAR equals the probability mass of the edit-distance distribution at zero. The UAR and edit depth therefore describe the two parts of one distribution and cannot contradict each other. Second, because the Levenshtein distance between two strings never exceeds the length of the longer string,^35^ δi lies between 0 and 1. Third, availability latency is an upper bound on reading time, so the latency arm supports one-sided inference only, and the UFA is the only measure in the family that can indicate approval without reading. Fourth, the family is defined only over content that passes through an approval. The inventoried actions that reach a patient or the record with no person accepting lie outside the measurement surface by construction. A UAR, however favorable, says nothing about them, which is why coverage is reported with it.

### Instrument status at the study commit

**Table 3 gives, for each route, what the system records at commit 687f45e295 and which measures it supports. Clinical documentation is the one route on which the family, apart from dwell latency and disposition, is** computable. The drafted and signed texts are both kept, the signer is recorded with a signature record, the commit time is the signature time, and availability can be taken from the finish time of the run that produced the draft, because documents carry no creation time. The model and prompt version are recorded on that run, which is linked to the document by encounter and agent. Documentation accept events were not written at all before a code change committed on November 16, 2026, because the writer referred to columns the table did not have. Until then the edit ratio was kept only on the document itself. Discards are not captured on this route. An unsigned draft is deleted when a newer draft of the same document is written (HZ-09).

On the help desk, edit telemetry and accept events are written when a draft is sent, but only if the review screen has stored the agent’s version of the draft. A draft approved without that stored version produces no telemetry row. The UAR and edit depth are therefore computable only on a captured subset, and acceptance coverage states its size. Attribution is incomplete by construction. The approver field on help-desk drafts is optional, the help-desk approve path records no user identifier (HZ-34), accept events can carry the label “an unattributed operator”, and the administration console accepts approvals without a named staff session unless a configuration setting requires one (HZ-33). Rejected and expired drafts are recorded.

On the WhatsApp agent route, the approve path records neither an edit signal nor an outcome, and, in the words of the hazard log, “a swept draft approved unchanged produces no telemetry row at all” (HZ-65). Whether an approval was edited cannot be established, so the UAR, edit depth and UFA are not computable on this route.

Availability latency can be derived from draft creation and approval times, rejected and superseded drafts are recorded, and attribution completeness can be measured. The approver is held as free text, which need not name a person, and drafts can be approved from a Slack card outside the review screen.

Other instruments are missing on every route or where they are needed. No route records when a draft is first opened for review, so dwell latency cannot be computed anywhere. No route has dedicated approval-latency capture, so availability latency is derived from creation and commit times that were written for other purposes. Draft records on the help-desk and WhatsApp agent routes carry no model or prompt identifier, so model stratification is possible only for clinical documentation. The operations-agent cards have no edit capture and no accept-event writer.

### Coverage at inventory level

Inventory coverage is the headline figure of inventory version 0.2, 29 of 153 actions taking effect without acceptance, with the reconciliation caveat given in Methods. Because the inventory records reach only for unaccepted actions, the 124 remaining actions cannot be divided from it into those that cross an approval boundary and those with no external effect. The analysis plan therefore reports inventory coverage from an itemized list of unaccepted actions, fixed before extraction, rather than from the headline count.

## Discussion

In this methods study at one private outpatient clinic, we specified a family of review-fidelity measures for LLM-drafted clinical content, defined at the event where a person commits a draft, and checked each against what the deployed system records. The measures are mutually consistent by construction, and the latency arm supports inference in one direction only. The check changed the paper. Only clinical documentation can support the family as specified. The help desk can support it on a subset whose size is itself a measure. The WhatsApp agent route cannot yet show whether an approval was edited, and no route records when a draft is opened.

The closest prior work analyzes edits after the fact. Guo and colleagues used a diff algorithm on 23,760 ambient notes and found that most were edited,^9^ which corresponds to an unedited share of about 15.6% at note level, and the same group showed that edits concentrate on clinical content.^10^ Jeanselme and colleagues modeled adoption, use and habituation separately and found that adoption and use were associated with turnaround time in opposite directions, with no further association after habituation over their study period.^13^ The components of the UAR family are not new. Edit rates, draft-to-final similarity and response time have been measured before,^9,11,12^ automated text metrics have been related to clinicians’ use of drafts,^15^ and the MHRA named review time and edit depth as candidate indicators.^24^ The family differs in four respects. It is defined on the approval event, it is intended to be computed continuously by the deployment rather than retrospectively over a study period, it is designed to sit within the deployment’s safety file, and it carries pre-specified rules for recognizing its own failure. Note-quality instruments such as the PDQI-9,39,40 the PDSQI-9 for LLM-generated summaries^41^ and harm-potential ratings of unedited output^17,42^ answer a different question. They measure what a draft or note contains and need expert raters, so they cannot run on every approval. They are the natural reference standard for the calibration audit.

The joint reading matters because acceptance is a weak signal on its own. If acceptance of incorrect recommendations runs close to acceptance of correct ones,^22^ and if trained physicians remain susceptible to erroneous suggestions,^21^ then a falling edit rate after a model update is uninterpretable without latency and an independent check of content. The unedited fast-approval rate combines the two candidate indicators named by the MHRA^24^ into one quantity computable on every approval where both texts and both times are kept. It cannot distinguish a slow but inattentive approval from a careful one, and only the calibration audit can estimate how many errors pass through each stratum.

The instrument check is itself a finding for other deployers. A system can be designed around human approval and still not record the approval in a form that supports measurement. In this deployment the gaps were of three kinds: the edited or unedited state of the draft was not kept, the person who approved was not named, and the time the draft was opened was never recorded. Each gap was invisible at the level of design documents and visible only in the code. Deployers who cite human review as a control could test their own systems against the five elements of the measurement surface before relying on any acceptance figure.

Three implications follow, each conditional on validation. For regulators and deployers, review-fidelity measures could serve as monitored controls within DCB0160 deployment safety cases and the post-deployment monitoring that NHS England guidance expects of deployers,^1,32^ and could support the automation-bias awareness that the EU AI Act requires of human oversight, whose obligations for AI in medical devices now apply from August 2, 2028.2,43 For vendors, a report of unedited acceptance^26^ would be more informative if accompanied by acceptance coverage, attribution completeness, latency and a calibration result. For evaluators, the instrument-failure rules suggest that implausible review-fidelity values should be investigated as defects in capture before they are interpreted as behavior.

### Limitations

This study has limitations. First, and most important, the measures have not been validated against missed errors. No evidence yet shows that a higher UFA predicts more errors reaching patients, and until the calibration audit reports, the family measures a necessary surface of review rather than review itself. Second, the setting is a single private outpatient clinic with a small team. The authors are describing their own system. Two authors hold equity in the developer and operator of the system, and one is the clinic’s clinical safety officer, appointed on October 18, 2026. The hazard-log version that matches the inventory analyzed here (version 0.2) was not approved, and no clinical safety case report has been written. NHS England guidance expects deployer monitoring to be independent of the manufacturer,^1^ and these measures would be computed by the operator. Their credibility depends on the integrity of the underlying records, which is examined in a companion manuscript.^44^ Third, three of the four routes cannot yet support the full family. On the WhatsApp agent route no review-fidelity measure beyond latency, attribution and disposition is computable, on the help desk the measures cover only a captured subset, and model stratification is possible only for clinical documentation. Fourth, attribution is incomplete on the messaging routes. A qualified and an unqualified approver produce the same UAR, and seniority of the approver is recorded but not checked (HZ-35), so the measures say nothing about whether the right person approved. Fifth, character-level distance is blind to meaning. A one-word change, such as an inserted negation, may be clinically decisive yet produce a small δ, so edit depth should be paired with edit categories of the kind developed in prior work.^10,11^ Sixth, coverage is incomplete in a second sense. Model-generated content, including live voice-agent speech, reaches patients without approval, and the inventory’s headline count of such actions does not reconcile exactly with its own list.

Seventh, staff who know their approvals are measured may change behavior, and a small team makes individual inference a risk that the disclosure rules reduce but cannot remove. Finally, the reading floor rests on general reading-rate evidence^36^ rather than on clinical review of drafts and will need empirical calibration, and for short drafts it may be shorter than the time needed to operate the review interface, which leaves the UFA insensitive on those drafts.

Whether these measures can be captured reliably once the missing instruments exist, and whether they detect review that fails to catch errors, are the questions that the first measurement window and the calibration audit are designed to answer. Multisite replication would be needed before they are used as general benchmarks.

## Conclusion

The UAR, with acceptance coverage, attribution completeness, edit depth, availability latency and the unedited fast-approval rate, makes the fidelity of human review of LLM drafts specifiable as telemetry at the approval event, without exporting patient content. In the deployment studied, the instruments support the full family on clinical documentation only. Deployments that rely on human review as a safety control could check that they record the approval event before reporting acceptance, and could report these measures jointly and with their coverage stated. Validation against missed errors is required before they are used to judge review quality.

## Supporting information

TRIPOD-LLM checklist

OSF registration - analysis plan

## Declarations

### Author contributions

All three authors contributed to the conceptualization of the work, drafted and reviewed the manuscript, and approved the final version, and all accept accountability for all aspects of the work. OE conceived the metric family and the study and drafted the manuscript. SY designed and implemented the capture instrumentation and directed the check of mechanism-level statements against the code. JD provided independent clinical safety review and critically revised the manuscript.

### Ethics statement

This methods article analyzed the system’s source code and configuration, its safety documents (the action inventory and hazard log) and the structure of its database, read with queries that returned no rows. No human participant was recruited, observed or surveyed, no patient or staff record was read, and no identifiable personal data were analyzed. The authors therefore did not seek research ethics committee review, and no committee reviewed this classification. No measure is computed or reported for any individual member of staff, and no output of this work feeds performance management. The analysis of window data, which concerns staff review behavior in aggregate, is not reported here. Patient and public involvement. Patients and the public were not involved in the design or reporting of this methods article.

### Data availability

No data were generated or analyzed beyond the code, configuration and documents described.

The metric definitions, interpretation framework and analysis plan are given in full in the text, the tables and Supplementary File 2. Drafted and committed text is clinical data and will not be shared. The code commit, inventory and hazard-log versions cited are available to editors and reviewers under confidentiality on request to the corresponding author.

### Code availability

The edit-telemetry code and, once written, the version-controlled extraction queries will be available to editors and reviewers under confidentiality on request. No proprietary source code is disclosed.

### Use of generative AI

The authors used large language model tools, including Claude (Anthropic), for drafting, editing and reference checking. On November 19, 2026, Claude (Anthropic; Claude Opus 5.5) was used, under the authors’ direction, to check the manuscript against the system’s code and database and to revise it. The software described was itself written with AI coding assistants (Claude models through Claude Code, and Cursor), as recorded in the repository’s commit history. The authors reviewed and verified all content and take full responsibility for the manuscript.

## Acknowledgments

The authors thank the clinical and engineering teams of the deployment.

