## Supplementary material for "The unedited acceptance rate: a review-fidelity metric for human-in-the-loop clinical AI": TRIPOD-LLM checklist

Supplementary File 1. TRIPOD-LLM checklist

**Manuscript.** The unedited acceptance rate: a review-fidelity metric for human-in-the-loop clinical AI (Elalfy, Youssef, Davids). Methods article with a pre-specified analysis plan.

**Guideline.** Gallifant J, Afshar M, Ameen S, et al. The TRIPOD-LLM reporting guideline for studies using large language models. Nat Med. 2025;31(1):60-69. Items and applicability codes are taken from the guideline's main checklist table (author manuscript, PubMed Central PMC12104976, read November 19, 2026). Item wording below is shortened.

**Research design and task.** The study defines measures of human review of LLM output in a health care setting, so items applicable to "LLM evaluation in healthcare settings" (H, with E and EH items) are answered. Items applicable only to de novo development or methods of model building (M, D, MD) are marked not applicable. The LLM tasks reviewed are documentation generation and drafted replies (conversational and long-form text). The article reports no measured values, so items that require results are marked "not applicable to a methods version" and are addressed in the analysis plan where possible.

Locations refer to section headings of the manuscript. Supplementary File 2 is the analysis plan in OSF Preregistration fields.

| **Item** | **Topic (shortened)** | **Applies** | **Where reported, or why not applicable** |
| --- | --- | --- | --- |
| 1 | Title identifies the study as evaluating an LLM, with task, population and outcome | All | Title; the article type line on the title page states a methods article with a pre-specified analysis plan |
| 2 | Abstract (TRIPOD-LLM for Abstracts) | All | Abstract, structured; see items 2a to 2l below |
| 3a | Health care context, use case and rationale, with existing approaches | All | Background and Significance, paragraphs 1 to 4 |
| 3b | Target population, intended use and intended users in the care pathway | EH | Materials and Methods, Setting and system (four approval routes, staff approvers, patients and the record as recipients) |
| 4 | Study objectives, including development, tuning or validation stage | All | Objective |
| 5a | Data sources for training, tuning and evaluation | All | No dataset was used. The study uses code, configuration, safety documents and database structure (Design and reporting; Ethics statement). Window data sources are specified in Supplementary File 2, Measured variables |
| 5b | Description of data points and their distribution | All | Not applicable to a methods version; data points per route are listed in Table 3 and Supplementary File 2, Measured variables |
| 5c | Dates of oldest and newest text items | All | No text items were analyzed. The window is defined in Pre-specified analysis plan, Window, and Supplementary File 2, Existing data |
| 5d | Pre-processing and quality checking | All | Metric definitions, Normalization; instrument-failure conditions in Pre-specified analysis plan |
| 5e | Handling of missing and imbalanced data | All | Pre-specified analysis plan (completeness rule, disclosure control); Supplementary File 2, Missing data and Data exclusion |
| 6a | LLM name, version and last date of training | All | Table 4 (model chains as configured on November 19, 2026). Training cut-off dates are not reported, as stated under Table 4 |
| 6b | LLM development process | MD | Not applicable; no model was developed or fine-tuned |
| 6c | How text was generated, prompts and inference settings | MDE | Partly applicable. Table 4 notes the reasoning setting; prompts are not reported because the study measures review of drafts, not generation |
| 6d | Initial and post-processed output of the LLM | All | Metric definitions (drafted text D and committed text C) and Normalization |
| 6e | Classification thresholds | All (COF tasks) | Not applicable; no classification task |
| 7a | Metrics of generative output quality | All | Metric definitions and Table 1. The measures describe review of output; output quality is the reference standard of the calibration audit (Proposed governance uses) |
| 7b | Relevance of metrics to the downstream task and correlation with human evaluation | EH | Interpretation framework, Table 2; Discussion, paragraph 3; Limitations, first item (no validation yet) |
| 7c | Outcome definition, formula and date of inference | EH | Metric definitions (formulas), Table 1; dates of inference will be the window dates (Supplementary File 2) |
| 7d | Qualifications of assessors for subjective assessment | All | Not applicable to this version; the calibration audit design will be registered separately (Proposed governance uses) |
| 7e | Comparison with other LLMs, humans or benchmarks | All | Not applicable; no comparison is made. Prior measures are compared in Discussion, paragraph 2 |
| 8a to 8c | Annotation | All | Not applicable; no annotation was done |
| 9a, 9b | Prompt design and data used for prompts | All | Not applicable; the study did not prompt LLMs |
| 10 | Pre-processing before summarization | All (SS tasks) | Not applicable |
| 11 | Instruction tuning or alignment | MD | Not applicable |
| 12 | Compute | MDE | Not applicable; the measures are computed by database queries on routine records |
| 13 | Ethics committee approval and consent | All | Ethics statement. No research ethics committee review was sought and no committee reviewed this classification |
| 14a | Funding and role of funders | All | Declarations, Funding |
| 14b | Conflicts of interest | All | Declarations, Competing interests |
| 14c | Where the protocol can be accessed | H | Supplementary File 2 (analysis plan in OSF Preregistration fields) |
| 14d | Registration information | H | Design and reporting: the plan will be registered on the OSF with a public time stamp before the window opens. It is not registered at the time of this version |
| 14e | Availability of study data | All | Declarations, Data availability |
| 14f | Availability of code | All | Declarations, Code availability |
| 15 | Patient and public involvement | H | Declarations, Patient and public involvement (none) |
| 16a to 16d | Participants and data flow | EH | Not applicable to a methods version; no patient or EHR data were analyzed |
| 17 | Performance on pre-specified metrics | All | Not applicable; no measured values are reported. The properties of the measures and the instrument status are in Results |
| 18 | LLM updating | All | Not applicable to this version; change points and model changes are handled in Pre-specified analysis plan, Instruments and change points |
| 19a | Overall interpretation | All | Discussion, paragraphs 1 to 4 |
| 19b | Limitations | All | Discussion, limitations paragraph |
| 19c | Known challenges in using data for the task | EH | Results, Instrument status at the study commit; Table 3 |
| 19d | Intended use, end-user and level of human oversight | EH | Setting and system; Proposed governance uses |
| 19e | Handling of poor-quality or unavailable input data | EH | Interpretation framework, fifth principle; instrument-failure conditions |
| 19f | User interaction and expertise required | EH | Setting and system (approval by staff and by clinicians who sign documents); Limitations, fourth item (approver seniority not checked) |
| 19g | Next steps for future research | All | Discussion, final paragraph |

**TRIPOD-LLM for Abstracts**

| **Item** | **Topic (shortened)** | **Applies** | **Where reported** |
| --- | --- | --- | --- |
| 2a | Title identifies the study | All | Title |
| 2b | Health care context and rationale | EH | Abstract, Objective |
| 2c | Study objectives | All | Abstract, Objective |
| 2d | Key elements of the setting | All | Abstract, Materials and Methods |
| 2e | Data used, splits and selective use | MDE | Not applicable; no data analyzed |
| 2f | Name and version of LLMs | All | Not given in the abstract for length; Table 4 |
| 2g | LLM-building steps | MD | Not applicable |
| 2h | Tasks performed by the LLMs | All | Abstract, Materials and Methods (drafting reviewed on four approval routes) |
| 2i | Evaluation datasets and endpoints | All | Abstract, Materials and Methods (measures); no dataset |
| 2j | Overall results and interpretation | All | Abstract, Results and Discussion |
| 2k | Broader implications | All | Abstract, Conclusion |
| 2l | Registration | H | Not in the abstract; the plan is not yet registered (Design and reporting) |
