## Supplementary material for "The unedited acceptance rate: a review-fidelity metric for human-in-the-loop clinical AI": OSF registration - analysis plan

### OSF registration content: analysis plan for the unedited acceptance rate

Prepared on November 19, 2026, for entry into the Open Science Framework (OSF) Preregistration template. Each heading below is one field of that template, in the template's order, with the text to paste. This document is also Supplementary File 2 of the medRxiv methods article. It is not a registration until it has been submitted on the OSF and has a public time stamp, and nothing in it should be read as saying that it has been registered.

Registration settings: template "OSF Preregistration"; no embargo, so that the time stamp is public at once; contributors Omar Elalfy, Sofian Youssef, Joseph Davids.

---

#### Study Information

##### Title

The unedited acceptance rate: a review-fidelity metric for human-in-the-loop clinical AI. Analysis plan for the first prospective 12-week measurement window.

##### Authors

Omar Elalfy (Wai Health Inc, Palo Alto, CA, USA; The London Wellness Clinic Ltd, London, UK), ORCID 0000-0001-7149-371X. Sofian Youssef (Wai Health Inc), ORCID 0000-0001-8494-4495. Joseph Davids (Department of Surgery and Cancer, Imperial College London; Department of Simulation, Imperial College Healthcare NHS Trust; Department of Neurosurgery, National Hospital for Neurology and Neurosurgery; London, UK), ORCID 0000-0002-9257-6722.

##### Description

Large language model (LLM) agents in a multi-agent clinical system (wai, developed and operated by Wai Health Inc) draft text that reaches patients or the clinical record at a private outpatient clinic in London (The

Wellness, operated by The London Wellness Clinic Ltd). Much of that text takes effect only when a person approves it. This plan fixes, before any of the data it governs exist, how the fidelity of that approval will be measured over a prospective 12-week window: acceptance coverage, attribution completeness, the unedited acceptance rate (UAR), edit depth, availability latency, the unedited fast-approval rate (UFA) and disposition.

The plan is written to what the system's instruments can measure, as checked against its code at commit 687f45e295 (November 19, 2026), its safety documents and read-only queries of database structure that returned no rows. It says plainly which instruments do not yet exist.

Instruments that exist at commit 687f45e295:

- Clinical documentation keeps both the drafted and the signed text, records the signer with a signature record, and records the signature time. The producing run records the model and prompt version and its finish time, and is linked to the document by encounter and agent. Accept events for documentation were not written before a code change committed on November 16, 2026.
- The help desk writes edit telemetry and accept events when a draft is sent, but only when the review screen has stored the agent's version of the draft. It records rejected and expired drafts, draft creation times and an approval entry in a separately written communications audit.
- The WhatsApp agent route records draft creation, approval and send times, rejected and superseded drafts, and an approver field held as free text.

Instruments that do not yet exist at commit 687f45e295:

- No route records when a draft is first opened for review. Dwell latency cannot be computed, and the plan uses availability latency only.
- No route has dedicated approval-latency capture. Availability latency is derived from creation and commit times written for other purposes.
- The WhatsApp agent route records no edit signal, so whether an approval was edited cannot be established (hazard HZ-65). The UAR, edit depth and UFA are not computed on this route unless an instrument is deployed and recorded under the amendment rule below.
- Help-desk and WhatsApp draft records carry no model or prompt identifier, so model stratification is possible for clinical documentation only.

- The approver field on the help-desk and WhatsApp routes need not identify a person. The help-desk approve path records no user identifier (HZ-34), and approval without a named staff session is possible by default (HZ-33).
- Clinical documentation does not capture discards. An unsigned draft is deleted when a newer draft of the same document is written (HZ-09).
- The operations-agent approval cards in Slack offer approve or reject and have no edit capture and no accept-event writer.
- The system has no field that marks test or synthetic records.
- The extraction queries have not been written. They will be committed, and their identifiers recorded in an amendment, before extraction.
- The calibration audit that would validate the measures against missed errors has not been designed. Its design will be registered separately before its first sample is drawn. It is not part of this registration.

Model-generated content also reaches patients or the record without any approval, for example live voice-agent speech and a live consultation recap pushed to the patient's screen. Such content is outside every measure here and is counted only as inventory coverage.

##### Hypotheses

None. This is a descriptive measurement study. No hypothesis is tested and no directional prediction is made. Any trend statement will be labeled exploratory.

---

#### Design Plan

##### Study type

Observational study.

##### Blinding

No blinding is involved in this study.

##### Is there any additional blinding in this study?

No analyst will view any drafted or committed text, approver identity or patient identifier. Distances, latencies, reading floors and counts are computed inside the clinical database, and only aggregates by route, stratum and week are exported.

##### Study design

Prospective observational measurement over a fixed 12-week window of routinely recorded approval events on three approval routes: the help desk (drafted replies to patient messages on several channels, including email and in-app messages), the WhatsApp agent route (drafts approved in the administration console or from a Slack approval card) and clinical documentation (drafted documents that a clinician signs). A fourth route, operations-agent approval cards, is described but has no measure other than inventory coverage. No intervention is made. Staff who approve drafts will be told of the measures and their purpose before the window opens, and the date will be recorded in an amendment.

##### Randomization

Not applicable.

---

#### Sampling Plan

##### Existing data

Registration prior to creation of data. The window opens at 00:00 Europe/London on the first Monday at least 14 days after the public time stamp of this registration, so none of the data it covers exist when the plan is registered.

##### Explanation of existing data

Approval records from before the window exist in the system but are not analyzed. The checks on which this plan rests read code, configuration, safety documents and database structure, with queries that returned no rows.

In November 2026, for companion manuscripts, the authors also saw aggregate counts of approver-field categories and of approval records created before September 21, 2026, from read-only queries that selected no text. Those records predate the window, are not part of this analysis and informed only the description of the instruments above.

##### Data collection procedures

No data are collected for the study. The system records approval events in the course of routine care. After the window closes, version-controlled extraction queries will run read-only against the clinical database. For each route they will compute, inside the database, the normalized texts, edit distances, word counts, reading floors and latencies of eligible events, and export only aggregate counts, proportions and quantiles by route, stratum and week. The query versions and extraction times will be recorded with each aggregate.

##### Sample size

All eligible approval events whose commit time falls in the window, on the help-desk, WhatsApp agent and clinical documentation routes. The number is not known in advance. Events cluster within approvers and weeks (12 week clusters).

##### Sample size rationale

No sample size calculation applies to a descriptive first window. The window length is fixed at 12 weeks. For orientation, the half-width of a 95% Wilson interval for a proportion near 0.5 is about 0.20 at 20 events, 0.10 at 96 events and 0.05 at 384 events.

##### Stopping rule

Data inclusion ends at 23:59:59 Europe/London on the Sunday that closes the twelfth week.

---

#### Variables

##### Manipulated variables

None.

#### Measured variables

For each approval event *i* on route *r*:

- Drafted text $D_i$ and committed text $C_i$, or a stored unedited flag. Clinical documentation: the drafted and signed document bodies. Help desk: the agent's stored version and the approved text, taken before translation or channel formatting. WhatsApp agent route: not available (see Description).
- Approver field, classified as a named person, blank, the name of an approval surface, or a generic label.
- Availability time $t_{avail,i}$: draft creation time (help desk, WhatsApp agent route); finish time of the producing run (clinical documentation, whose documents carry no creation time).
- Commit time $t_{commit,i}$: approval entry in the communications audit (help desk); approval or send time (WhatsApp agent route); signature time (clinical documentation).
- Disposition: rejected or expired (help desk); rejected or superseded (WhatsApp agent route).
- Draft type, route, and, for clinical documentation, the model and prompt version recorded on the producing run.
- Commit record for coverage: approval entries in the communications audit (help desk); drafts reaching approved, sending or sent status (WhatsApp agent route); signed documents with their signature record and audit-chain event (clinical documentation).

#### Indices

- Normalization *N*: collapse every run of whitespace, including line breaks, to one space and trim both ends; for clinical documentation, first remove markup tags and non-breaking-space entities. No Unicode normalization is applied and no appended element is removed, because the committed text is taken at the approval step. This is the normalization the system's edit telemetry applies.
- Unedited: $N(C_i) = N(D_i)$.
- Edit distance $\delta_i = \mathrm{lev}(N(D_i), N(C_i)) / \max(|N(D_i)|, |N(C_i)|)$, with Levenshtein distance and length counted in UTF-16 code units, as implemented in the system. $\delta_i$ lies between 0 and 1, and $\delta_i = 0$ exactly when the event is unedited. Sensitivity analysis: the same ratio over whitespace-delimited tokens.

- Unedited acceptance rate (UAR): share of complete events that are unedited.
- Edit depth: distribution of $\delta_i$ among complete events with $\delta_i > 0$.
- Availability latency $L_i = t_{commit,i} - t_{avail,i}$, in seconds. It is an upper bound on reading time.
- Reading floor $f_i = 60 \cdot n_i / 300$ seconds, where $n_i$ is the whitespace-delimited word count of $N(D_i)$. Sensitivity analysis: 238 words per minute.
- Unedited fast-approval rate (UFA): among complete events with a latency value, the share that are unedited with $L_i < f_i$. Reported with the conditional share of unedited events that are fast.
- Acceptance coverage: share of commits in the route's commit record that are matched to an approval event with complete capture.
- Attribution completeness: share of approval events whose approver field identifies a named person.
- Disposition: share of drafts rejected, expired or superseded without commit, where captured.
- Inventory coverage: number of inventoried actions that take effect with no person accepting, from an itemized list of the action inventory fixed and recorded in an amendment before extraction.

---

### Analysis Plan

#### Statistical models

Descriptive only. For each route and week: acceptance coverage, attribution completeness, UAR, UFA and the conditional fast share as counts and proportions with Wilson score 95% confidence intervals; edit depth as the median, 75th and 90th percentiles of δ among edited events; availability latency as the median and 90th percentile; disposition as a proportion. Window-level estimates are given with Wilson intervals and, as a sensitivity analysis, a cluster bootstrap that resamples weeks (2,000 replicates). With 12 week clusters and few approvers, neither interval fully reflects clustering by approver, and window-level intervals are interpreted as approximate. Weekly series are shown graphically.

Planned output table, by route: approval events (n), acceptance coverage (%), attribution completeness (%), UAR (% and 95% CI), edit depth (median and 90th percentile), availability latency (median and 90th percentile, seconds), UFA (% and 95% CI) and number of instrument-failure weeks out of 12, with a final row for all routes reporting counts only, as composition.

#### Transformations

Text normalization as defined under Indices. Draft word count is divided into tertiles within each route, with cut points computed over the whole window. Approver fields are recoded into four categories (named person, blank, approval-surface name, generic label) inside the database.

#### Inference criteria

No hypothesis tests and no significance thresholds. Estimates are reported with 95% confidence intervals and interpreted descriptively under the joint-reading rules of the methods article (Table 2 there): no single measure is read alone, latency is informative in one direction only, readings are stratified, and instrument failure is ruled out before behavior is interpreted.

#### Data exclusion

- Events on records in a list of test records fixed before the window opens. The system has no field that marks test records, so the list is the only exclusion rule; its size, but not its content, will be recorded in an amendment.
- The operations-agent route, from every measure except inventory coverage.
- Incomplete events (no drafted text, no committed text or stored unedited flag, no approver field or no commit time) from the UAR and edit depth. They are counted and reported as a completeness proportion by route and week.
- Instrument-failure route-weeks from behavioral interpretation. A route-week is an instrument failure if (1) acceptance coverage is below 95%; (2) completeness among captured events is below 95%; (3) the UAR is exactly 0 or 1 with at least 20 events, unless both capture arms are shown to work by writing edited and unedited approvals on synthetic records in a non-production environment; or (4) for clinical documentation, more than 5% of events cannot be linked to their producing run. Failed route-weeks are counted and reported as instrument findings.
- Measures on any route before the deployment date of the instrument they depend on, as recorded under the amendment rule.

#### Missing data

Missing latency is reported separately, and the UFA is computed only on events with a latency value. No imputation is used. Events without a named approver are kept in all counts but cannot count toward distinct approvers, so they are reported only at route level over the whole window.

Disclosure control: any cell with fewer than five events is suppressed; wherever one cell in a row or column is suppressed, at least one further cell in it is suppressed so that no suppressed value can be derived by subtraction; any cell whose approvals come from fewer than three distinct named approvers is reported only after aggregation over a longer period or across strata; counts by approver are never reported.

##### Exploratory analysis

Trend over the 12 weeks, comparison across model or prompt versions on clinical documentation, and comparison across change points. All will be labeled exploratory.

---

#### Other

Amendment rule. Before the window opens, one dated amendment will record: the date staff were told of the measures; the model configuration in force (the models configured on November 19, 2026, are listed in Table 4 of the methods article); the deployment date of any change to the instruments listed above made after commit 687f45e295; and the size of the test-record list. Before extraction, a second dated amendment will record the extraction query identifiers, the itemized list of unaccepted actions used for inventory coverage, and any change point during the window. A change to an instrument or a model during the window is a change point, and estimates are reported separately on either side of it. Deviations from this plan will be reported with reasons.

Governance. No measure is computed or reported for any individual, and no output feeds performance management. Proposed governance uses of the measures (threshold monitoring, change evaluation and a calibration audit) are described in the methods article and are not in operation.

Ethics. The analysis concerns staff review behavior in aggregate. The authors have not sought research ethics committee review for the methods article, and no committee has reviewed this plan. The results report will state the position taken for the window analysis.

Conflicts of interest. Omar Elalfy and Sofian Youssef hold equity in Wai Health Inc, which developed and operates the system, and are directors of The London Wellness Clinic Ltd, where it is deployed. Sofian Youssef is Chief Technology Officer and Chief Operating Officer of Wai Health Inc and the clinic's clinical safety officer. Joseph Davids declares no competing interests. Funding: Wai Health Inc internal funds; no external funder.

Related manuscript. The methods article "The unedited acceptance rate: a review-fidelity metric for human-in-the-loop clinical AI" (Elalfy, Youssef, Davids) gives the definitions, their properties, the instrument check and the interpretation framework.
